# The distribution of asymptomatic disease duration in transmission models for tuberculosis: implications for epidemic projections and the impact of active case finding

**DOI:** 10.64898/2026.10.21.26366009

**Authors:** Zhichao Zhou, Nicolas A Menzies, Ted Cohen

## Abstract

Tuberculosis (TB) transmission is sustained in part by asymptomatic disease(aTB). This limits the effectiveness of passive case finding interventions that rely on symptom-based diagnosis. Compartmental transmission models commonly represent the time spent with aTB as a single disease state, thereby imposing an exponentially distributed waiting time and a constant hazard of leaving that state. We evaluated how differences in the shape of the aTB waiting-time distribution affect the fit to observed disease progression data and subsequent projections of TB incidence and active case finding (ACF) impact. We fitted exponential and chained-state gamma formulations to cohort data on aTB progression and incorporated them into transmission models. We found the gamma-distributed model to fit the data at least as well the exponential model, indicating that the alternative model structures can also capture changes of aTB dynamics. Comparing the fitted gamma-distributed model and the exponential model, we quantify how assumptions about aTB dynamics affect future disease trajectories and the expected impact of ACF interventions. Using India as a case study, we found that the gamma-distributed model projects a slower decline in TB incidence under the status quo, but a substantially larger reduction in the projected incidence of TB when ACF is employed, compared with the exponential model. These results suggest that estimates of ACF impact, and consequently assessments of its cost-effectiveness, may be sensitive to assumptions regarding the shape of waiting-time distribution for asymptomatic disease, indicating the importance of better evidence on the natural history of asymptomatic disease for projecting of intervention effects.

**Significance Statement:** Compartmental tuberculosis (TB) models often assume constant transition rates between natural history states, leading to exponentially distributed waiting times. However, this assumption may not be realistic for all state transitions, and may have consequences for disease dynamics and projected responses of the system to interventions. Here, we use cohort data to evaluate alternative assumptions about the distribution of time spent with asymptomatic TB and incorporate these assumptions into a transmission model. We show that when the mean of waiting-time is fixed, its shape of the distribution can substantially influence the projected impact of active case-finding interventions, even when alternative distributions produce similar baseline incidence trajectories. These findings highlight the importance of carefully evaluating waiting-time assumptions and their associated structural uncertainty when using transmission models to estimate intervention effects.

## 1 Introduction

Population-based tuberculosis (TB) prevalence surveys conducted over the past three decades have revealed that a substantial fraction of individuals with prevalent, bacteriologically-positive TB do not report the presence of typical symptoms of TB, such as persistent cough or weight loss [1, 2]. Here, we refer to this disease state as “asymptomatic” TB (aTB, consistent with the most recent WHO guidance [3]), but terms such as “sub-clinical” or “early” have also been used [4, 5]. A meta-regression using data from all available surveys (23 national and 5 subnational surveys) concluded that 50.4% (interquartile range, IQR, 39.8%-62.3%) of all prevalent TB was asymptomatic [5].

The clinical and public health implications of the finding that approximately half of all prevalent bacteriologically-positive TB is found among individuals who do not report typical symptoms are potentially profound [5]. Because most tuberculosis control programs rely on passive case finding (i.e., individuals must self present to health services for diagnosis) [6], individuals are unlikely to be diagnosed or treated while in an asymptomatic disease state. This results in negative individual-level and community-level consequences: delayed diagnosis of individuals may lead to worse short- and long-term health outcomes [7] and may allow longer periods of disease transmission in communities. The magnitude of these negative health outcomes depends on both the natural history of aTB (i.e., how long does this disease state last, and how many progress to symptomatic disease or self-cure?) and the infectiousness of individuals with aTB [8]. Recent research has focused on both of these areas of uncertainty, providing evidence that those with aTB can self-cure or progress to symptomatic TB [9], but may also remain in this disease state for years [10–13]. Further, individuals with aTB can have smear-positive TB [14] and may generate as many secondary cases as individuals with symptomatic disease [15–17]. A recent modeling analysis estimated that more than two-thirds of transmission in Asia and Africa may be attributable to those with aTB [2, 16]. Transmission dynamic models designed to incorporate this growing understanding of the complex natural history of infection have typically included aTB as a single health state [9, 11, 18, 19], encoding the implicit assumption that there is a constant hazard of progression from this state and exponentially distributed durations of aTB. In this paper, we examine how the distribution of time individuals spend in an asymptomatic infectious state influences future incidence trends and the effectiveness of ACF interventions. First, we test whether models that allow for non-exponentially distributed durations of aTB provide a comparable fit to observed natural history data, by fitting a range of model structures to an empirical dataset on TB incidence rates for individuals with aTB. Second, we examine the consequences of incorporating these different structures into transmission models to project epidemic trajectories and to estimate the effect of case finding interventions. We use India as our motivating example, since India bears the world’s largest TB burden [20, 21]. We use epidemiological data, including national incidence estimates and findings from an Indian cohort study [22], to ground our assumptions and model parameters in real-world observations [19, 20, 22]. By explicitly linking assumptions about the duration and progression dynamics of aTB to epidemic projections and intervention impact, we find that the details of the natural history of infection have important consequences for epidemic projections and anticipated effects of interventions [2].

## 2 Methods

### 2.1 Model Structure

Figure 1 illustrates the structure of the model. Individuals progress from minimal disease to asymptomatic tuberculosis (aTB), and subsequently to symptomatic disease. Minimal disease is defined here as pathological evidence of disease prior to bacteriological confirmation of TB, regardless of the presence of symptoms [11, 23]. In the exponential model, aTB is represented as a single compartment. In the gamma-distributed formulation, the asymptomatic stage is divided into *k* sequential sub-states [24, 25].

**Figure 1:**
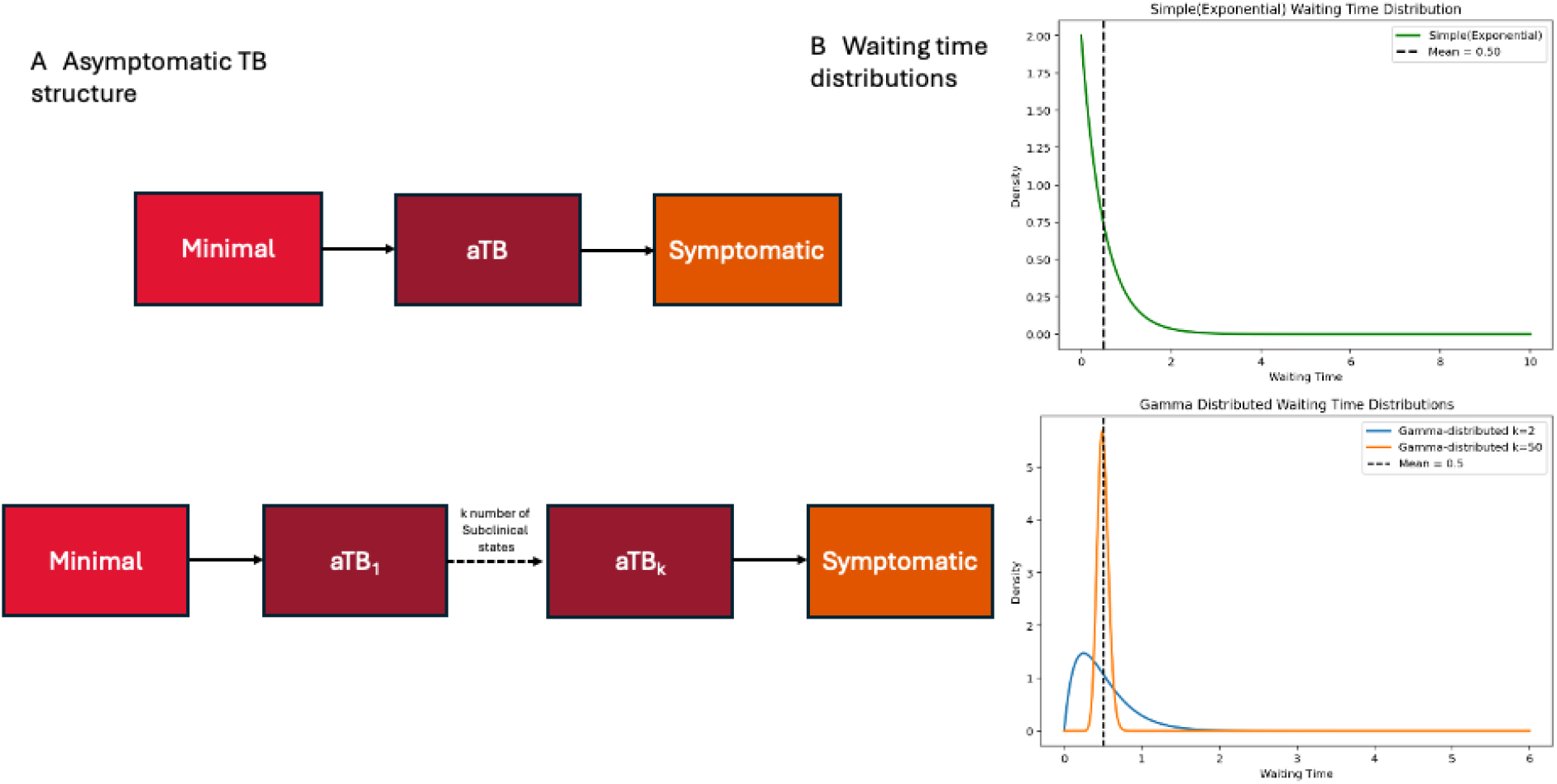
(A) Compartmental representation of asymptomatic TB progression. Individuals progress from minimal disease to aTB and subsequently to symptomatic disease. In the exponential model, aTB is represented as a single compartment. In the gamma-distributed formulation, the asymptomatic stage is divided into *k* sequential sub-states (aTB_1_, …, aTB_*k*_). (B) Waiting time distributions implied by the two model structures. The single-compartment formulation corresponds to an exponential waiting time distribution, whereas the multi-stage formulation produces an gamma distribution with the same mean (6 months) but reduced probability of extremely short durations.

Let *η* denote the progression rate from aTB to symptomatic disease in the exponential model. To preserve the same mean duration of the asymptomatic stage in the gamma-distributed formulation, each of the *k* sequential sub-states is assigned the same transition rate, *κ* = *kη*. The total time spent across the *k* sub-states therefore follows an gamma distribution with shape *k* and rate *κ*, yielding a mean duration of *k/κ* = 1*/η*, identical to that of the exponential model.

This parameterization ensures that the differences between the model outputs reflect the differences in the shape of the waiting time distribution rather than the differences in its mean. In particular, the gamma-distributed formulation relaxes the memoryless assumption of the exponential model and reduces the probability of extremely short asymptomatic durations.

### 2.2 Do cohort data support adoption of a model with non-exponentially distributed durations of aTB?

To evaluate whether the exponential distribution of aTB durations adopted in previous modeling studies adequately reflects empirical observations, we conducted a model-fitting analysis using cohort data that captured progression from aTB to symptomatic TB. We implemented a series of gamma-distributed models with increasing shape parameters (*k* = 1 (e.g., a exponential model assuming constant hazard) to *k*=100 (a model with a sharply peaked distribution of waiting times), holding the expected duration constant, to assess the performance of alternative gamma-distributed models. Model fits were compared using the widely applicable information criterion (WAIC) [26, 27] and leave-one-out information criterion (LOOIC) [28].

### 2.3 Does the distributional assumption of the duration of aTB affect projected TB trajectories?

To assess whether the distribution of aTB duration affects projected TB trends, we extended the natural history framework to a population-level transmission model. Two models were calibrated to match India’s TB epidemic in 2022: one assuming constant progression (i.e., an exponential model (*k* = 1)) and the other using the gamma-distributed structure (*k >* 1). We calibrated each model to WHO estimated TB incidence and mortality using a Bayesian framework [12, 29, 30]. We then projected TB incidence under the status quo from 2024 to 2033 and compared the two models’ predictions to examine differences in the trajectory of TB burden over time.

### 2.4 Does it matter for the estimated health impact of ACF interventions?

To assess whether assumptions about the distribution of aTB disease duration affect the estimated health impact of ACF, we conducted counterfactual intervention simulations under alternative natural history models. Here, ACF is conceptualized as an intervention that does not rely on symptom-based pre-screening, thereby allowing detection of aTB. We assume that individuals in the aTB state are not routinely identified through passive symptom-based healthcare seeking and, therefore, remain eligible for detection through ACF. ACF was implemented as a temporary increase during 2024–2026 in two state transitions: movement from aTB to minimal disease (reflecting earlier detection and entry into the interim recovery pathway) and from minimal disease to recovery (reflecting treatment initiation and completion). In the model, this was represented as a proportional increase in both transition rates over the intervention period. Again, we compared an exponential aTB duration model with the best fitting gamma-distributed model. For each model, an ACF intervention lasting three years was simulated and epidemic trajectories were compared to evaluate the sensitivity of the projected ACF impact to our distributional assumptions for the duration of aTB.

## 3 Results

### 3.1 Gamma-distributed models provide comparable fit to Indian cohort data

We first evaluated the ability of the 7 alternative model structures to reproduce subclinical progression patterns observed in the National Tuberculosis Institute (NTI) cohort study in Bangalore, India [19, 22, 31]. Models with varying numbers of aTB states (*k* = 1 to 100) were fit to the data and compared using the WAIC and LOOIC. As shown in the Table 1, introducing multiple aTB substates produced model fits that were broadly comparable to the Exponential model. Because the differences in WAIC and LOOIC between models were modest, these measures provided limited evidence to distinguish a specific value of *k*. We therefore selected the *k* = 50 formulation for subsequent analyses because it had the lowest WAIC and LOOIC among the models considered. It is interpreted as a plausible representative gamma-distributed specification rather than a uniquely supported model. Figure 2 further shows that the exponential and gamma-distributed models produced broadly similar fits to the observed data, with only modest differences between them.

**Table 1:**
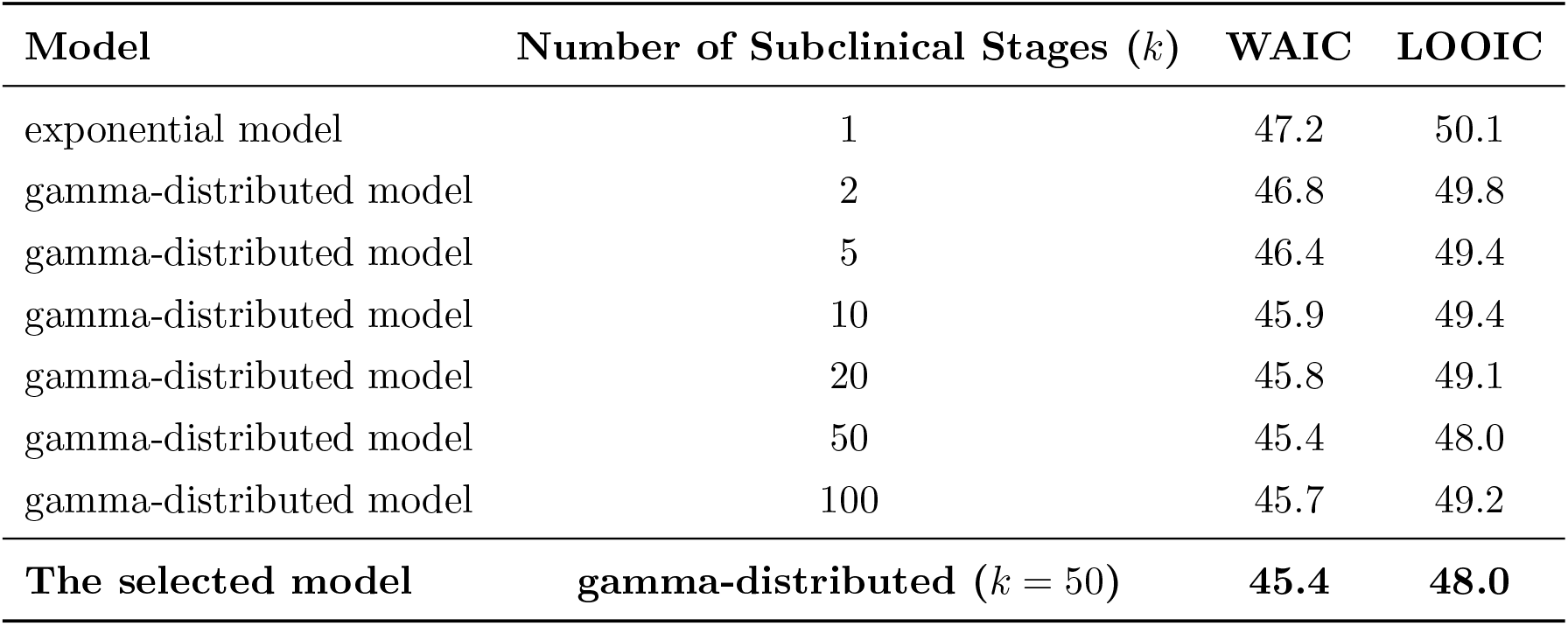
Model fit comparison using WAIC and LOOIC.

| Model | Number of Subclinical Stages ($k$) | WAIC | LOOIC |
| --- | --- | --- | --- |
| exponential model | 1 | 47.2 | 50.1 |
| gamma-distributed model | 2 | 46.8 | 49.8 |
| gamma-distributed model | 5 | 46.4 | 49.4 |
| gamma-distributed model | 10 | 45.9 | 49.4 |
| gamma-distributed model | 20 | 45.8 | 49.1 |
| gamma-distributed model | 50 | 45.4 | 48.0 |
| gamma-distributed model | 100 | 45.7 | 49.2 |
| **The selected model** | **gamma-distributed ($k = 50$)** | **45.4** | **48.0** |

**Figure 2:**
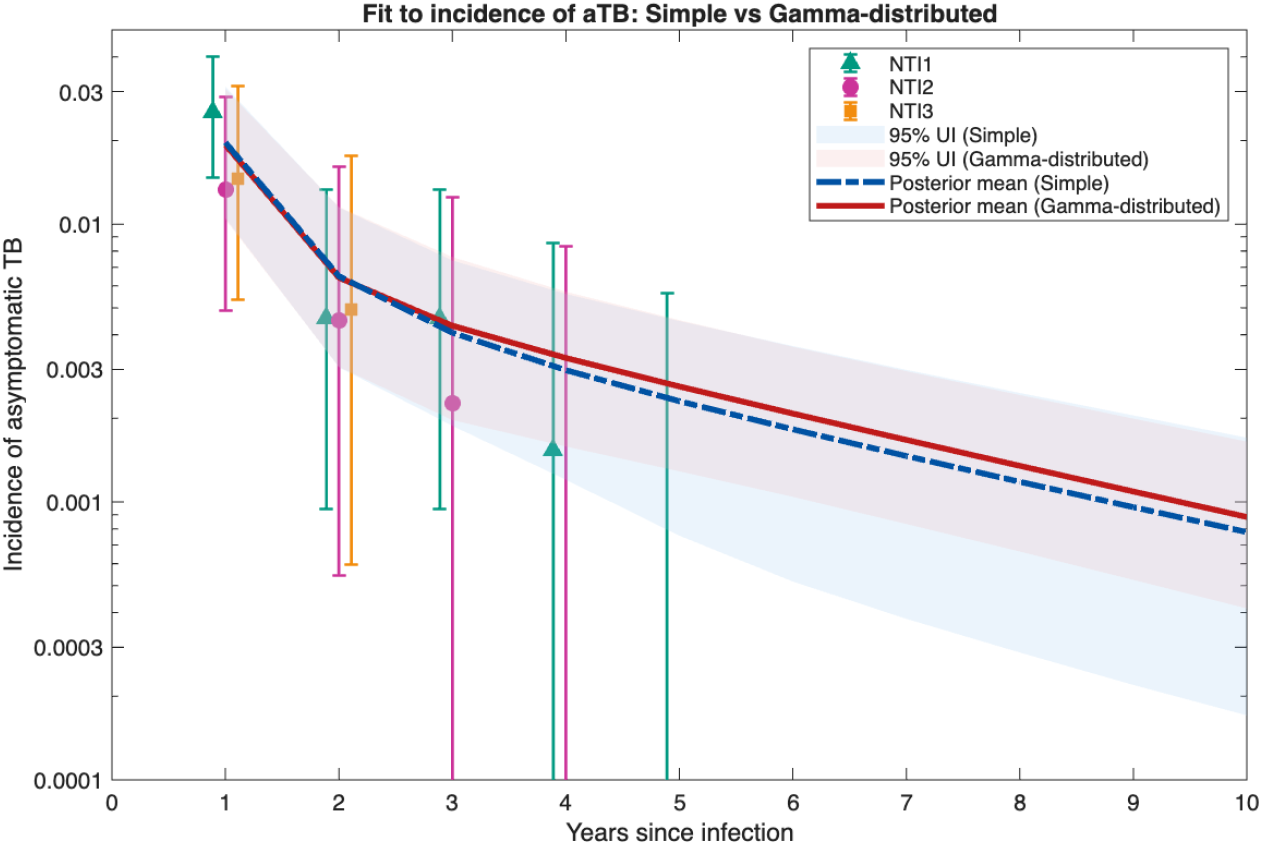
Comparison between exponential model and the selected gamma-distributed model (note log-scale for y-axis). Points labeled NTI1–NTI3 denote three data series derived from the National Tuberculosis Institute study((see Supplementary Information))

### 3.2 Gamma-distributed models of aTB project slower declines in TB incidence in India the absence of additional interventions

The results illustrate two modeled trajectories of TB incidence in India: one calibrated using an exponential model, one using an gamma-distributed model (Figure 3). Both model specifications are able to fit the observed temporal pattern of TB incidence in India as estimated by WHO [32] over the calibration period. The empirical time series show small positive deviations post-2020, likely reflecting changes in the TB epidemic attributable to COVID-19 [33], which we have not attempted to reproduce here. Forward projection of these fitted models show that the gamma-distributed( *k* =50) model produces slightly higher estimates of the future incidence of TB. By 2033, the projected TB incidence was 129.3 cases/100,000 (95% UI: 109.5-148.7) for the gamma-distributed model, 4.9% higher than for the exponential model (123.3 cases/100,000 (95% UI: 100.5-191.1)). These results suggest that assumptions regarding subclinical progression influence long-term disease projections.

**Figure 3:**
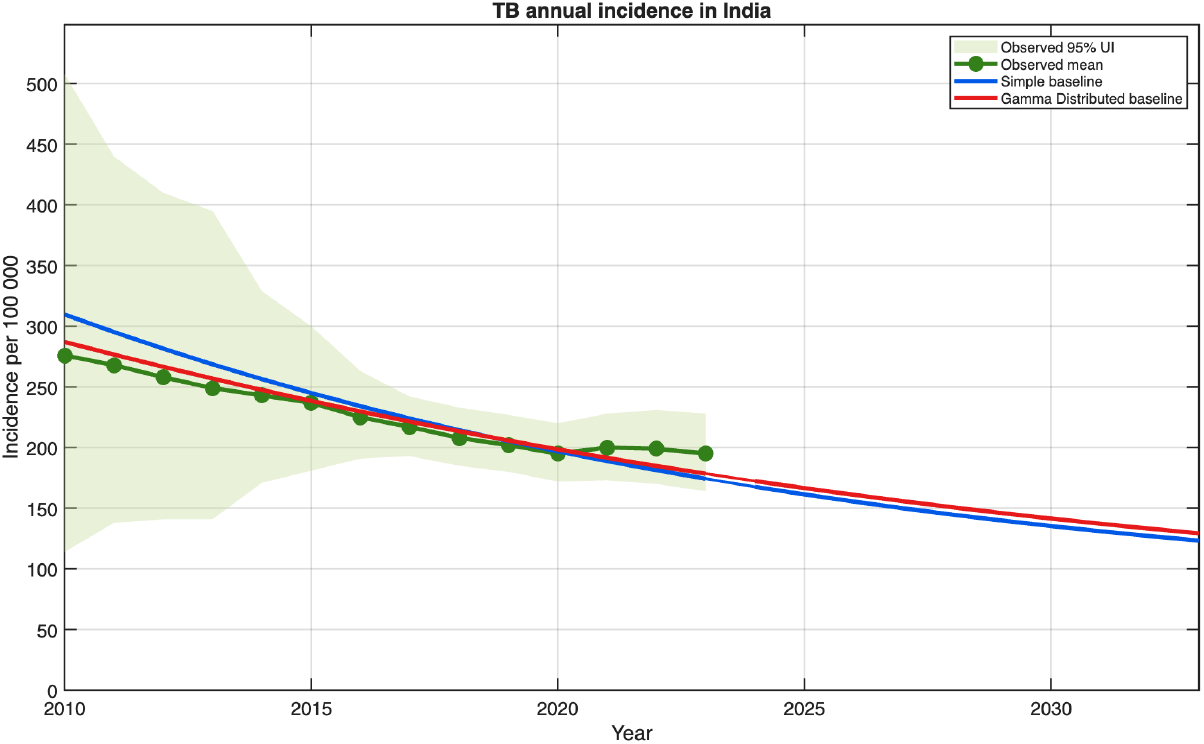
Projected TB incidence in India under alternative progression assumptions

### 3.3 ACF has greater projected impact on TB incidence reduction in gamma-distributed models of aTB

To examine the sensitivity of projected effects of ACF under the exponential and gamma-distributed models, we compared the effects of a three-year ACF intervention implemented from 2024 to 2026 (Figure 4a). For the exponential model, the expected TB incidence in 2026 (at the end of the ACF intervention) was 30.5 cases per 100,000 lower under ACF than under the no-intervention baseline (a reduction from 155.5 (95%UI 140.8, 186.4) to 125.0 (95%UI 104.3, 146.2)), representing a 19.6% reduction attributable to ACF. For the gamma-distributed(50) model, the expected impact of ACF was larger: 50.3 cases per 100,000 population (a reduction from 161.1 (95%UI 146.4, 175.7) to 110.8(95%UI 89.8, 128.0)), representing a 31.2% reduction attributable to ACF. Compared with the exponential model, the gamma-distributed model projects an ACF effect that is 59.2% larger in terms of relative TB incidence reduction at the end of this three-year intervention period.

**Figure 4:**
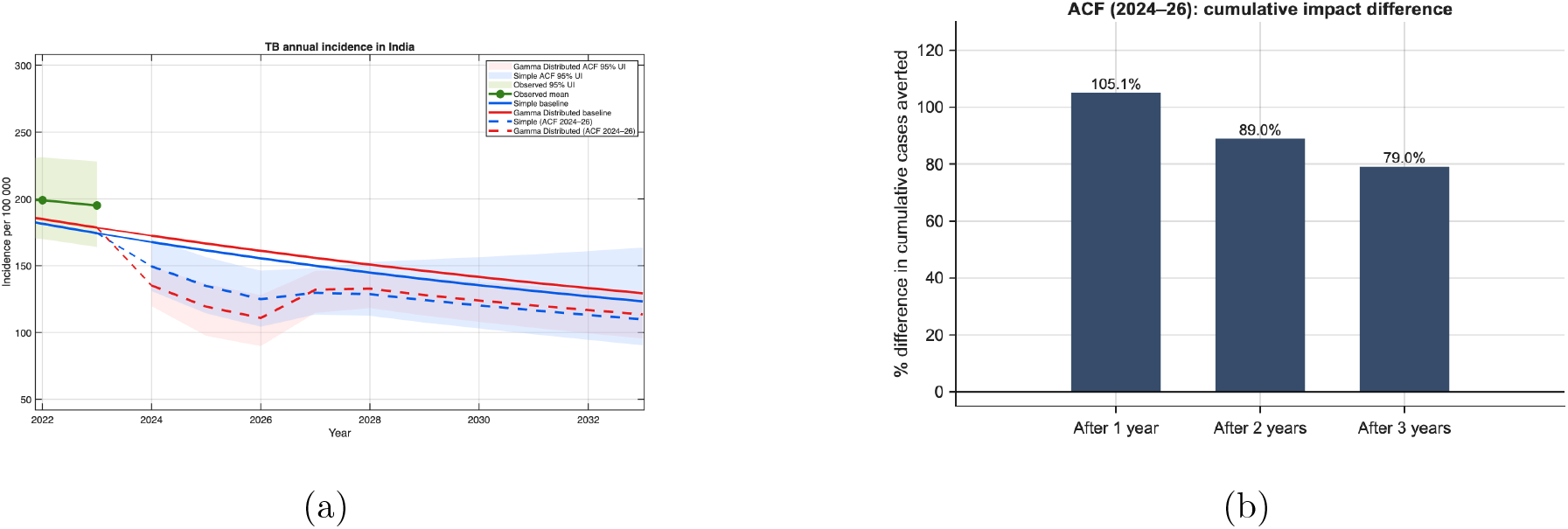
Comparison of baseline incidence projections and cumulative cases averted under ACF in India, contrasting the gamma-distributed and exponential progression models. (a) The left panel presents projected TB incidence trajectories under baseline conditions and under a three-year ACF intervention (2024–2026) for both progression assumptions. (b) The right panel shows the cumulative percentage difference in TB cases averted under ACF between the two models, calculated separately for each year of the intervention period (2024, 2025, and 2026). Cumulative cases averted under each model were calculated as the difference between baseline and ACF incidence, summed annually from 2024 onwards. The cumulative percentage difference between models was defined relative to the exponential model.

In 2033 (seven years after the end of the modeled ACF intervention), the incidence reductions produced by the ACF intervention, relative to the no-intervention baseline, were still evident but reduced in magnitude for both the exponential and gamma-distributed models. In 2033, the exponential model estimates an absolute reduction of 13.4 incident cases per 100,000 population (10.9%) attributable to ACF, whereas for the gamma-distributed model, the corresponding reduction is 15.8 incident cases per 100,000 population (12.2%).

The comparison of cumulative TB cases averted (Figure 4b) indicates differences in projected intervention impact between models. Compared with the exponential model, the gamma-distributed(*k* =50) model estimated approximately 2.05 times as many cumulative cases averted after the first year of intervention, corresponding to a 105.1% increase in impact. While the relative difference declines over time (an increase of 89.0% after two years and 79.0% after three years of ACF), the gamma-distributed model projects more cumulative cases averted than the exponential model throughout the intervention period.

### 3.4 Sensitivity of findings to assumptions of the impact of ACF

To assess the sensitivity to ACF strength, we varied the magnitude of increases in case detection (*m*). Larger values of *m* correspond to stronger effects on both detection and subsequent treatment initiation. Table 2 shows that the divergence in predicted impact between the gamma-distributed and exponential models increases as the ratio of TB cure achieved by the intervention increases, with the gamma-distributed model predicting progressively lower TB incidence than the exponential model.

**Table 2:** Structural difference between the gamma-distributed and exponential models under 3-year ACF (2024–2026).

| Multiplier $m$ | 2024 | 2025 | 2026 |
| --- | --- | --- | --- |
| 1.2 | -3.7 (-2.3%) | -3.9 (-2.6%) | -3.2 (-2.3%) |
| 1.5 | -14.3 (-9.6%) | -15.5 (-11.5%) | -14.2 (-11.3%) |
| 1.7 | -20.3 (-14.2%) | -21.8 (-17.3%) | -19.9 (-17.3%) |
| 2.0 | -27.7 (-20.5%) | -29.4 (-25.6%) | -26.6 (-26.0%) |

## 4 Discussion

Recent empirical and modeling work has emphasized the clinical and epidemiological importance of aTB. In this work, we investigated whether more complex models of progression from aTB than have typically been adopted are appropriate, and whether these models produce meaningful differences in disease projections and the expected impact of interventions. We evaluated assumptions governing the distribution of the duration of aTB by fitting alternative (i.e., exponential and gamma-distributed) model structures to individual-level data from the NTI Bangalore cohort. We then constructed and fit each transmission model to a common population-level baseline using national TB incidence estimates from WHO, enabling a comparison of how these alternative subclinical progression assumptions influence long-term projections and intervention effects.

We found that the gamma-distributed and exponential formulations provided broadly comparable fits to the observed progression data. In projections without ACF, the two model structures produced broadly similar incidence trajectories, although gamma-distributed model consistently projected modestly higher incidence than the exponential model. When projecting the impact of ACF interventions, differences between model structures were substantially more pronounced. Relative to their respective baselines, the gamma-distributed model predicted a larger percentage reduction in incidence than the exponential model, reflecting the sensitivity of case finding interventions to the distribution of waiting times in the aTB state.

In state-transition models for infectious diseases, assumptions about the distribution of time spent in each disease state are often made for mathematical convenience rather than empirical realism [34]. These models typically adopt a single compartment for each relevant natural history state, from which there is a constant hazard of transition and therefore exponentially-distributed waiting times. Others have demonstrated that even when expected durations of time spent in a state are held constant, the shape of the waiting-time distribution can substantially alter epidemic dynamics [35]. Non-exponential formulations, such as gamma-distributed or more general phase-type representations, introduce time-varying hazards and reduce the probability of early transitions. These different assumptions lead to different patterns of state occupancy, which have been shown to influence persistence and oscillatory behavior in epidemic systems [36], as well as the sensitivity of model outputs to parameter perturbations [37].

These considerations may be particularly important for TB, where long and heterogeneous disease courses may challenge the validity of constant-hazard assumptions [4]. Previous work has demonstrated that replacing commonly used exponential assumptions about demographic processes such as survivorship, with more realistic distributions can alter TB transmission dynamics and intervention effects [38]. Our findings are consistent with this broader insight, suggesting that assumptions about subclinical progression distributions may influence estimates of ACF intervention impact, which may have important implications for the cost-effectiveness of such strategies [39]. More broadly, these results highlight the importance of treating distributional assumptions as a source of uncertainty in state-transition models, especially when evaluating interventions that depend on the distribution of waiting times in disease states.

A central finding of this study is that relatively modest changes in model structure can lead to substantial differences in epidemic dynamics and projected intervention impacts over time. Although the two formulations share similar compartmental representations and identical mean asymptomatic disease durations, they differ fundamentally in how progression risk is distributed over time.

In the absence of intervention, baseline projections of incidence remain similar over shortl time horizons, but systematic differences emerge over longer periods. In the Indian context, the exponential model projects lower longer-term TB incidence relative to models with a more realistic temporal structure of aTB progression. While these differences are relatively small compared with the overall similarity in model fit and baseline projections, they illustrate how alternative assumptions regarding aTB progression can influence long-term epidemic forecasts.

While there are modest effects on long-term epidemic behavior, the most notable implication of the difference in the distribution of aTB waiting times is on the estimated impact of ACF interventions. Because an ACF intervention can allow for identification and and treatment individuals with aTB, its estimated effectiveness depends in part on how long individuals remain in the aTB state. Exponential models that suggest substantial rapid progression from aTB inherently limit the opportunity for earlier detection, while more complex models which concentrate the distribution of waiting times closer to the mean may provide more opportunity for detection of individuals in the aTB state.

These structural differences also have implications for cost-effectiveness analyses. Economic evaluations of detection-based interventions typically depend on projected cases averted and the scale of screening required. If simpler models systematically underestimate the epidemiological impact of ACF, they will likewise underestimate its cost-effectiveness, even when intervention costs are held constant. While we do not conduct a formal economic evaluation here, these findings suggest that structural assumptions about disease progression can influence not only epidemiological conclusions, but also policy-relevant economic assessments.

## Data Availability

All data produced are available online at GitHub

https://github.com/Lionel0923/tb-erlang-progression-model.git

## Data Availability

Replication data and analyses scripts are available on GitHub (https://github.com/Lionel0923/tb-erlang-progression-model.git).

## Acknowledgments

Research reported in this manuscript was supported by the National Institute of Allergy and Infectious Diseases of the National Institutes of Health under award number R01AI180209. The content is solely the responsibility of the authors and does not necessarily represent the official views of the National Institutes of Health.

## Author Contributions

ZZ and TC developed the original concept for this manuscript. ZZ developed the models and conducted the analyses. TC and NM reviewed analysis. ZZ wrote the first draft of the manuscript. TC and NM contributed to writing the final version of the paper. TC was responsible for supervision and funding.

