## Supplementary Information for "The distribution of asymptomatic disease duration in transmission models for tuberculosis: implications for epidemic projections and the impact of active case finding"

### Supplementary Material

#### Distributional Assumptions in Asymptomatic Tuberculosis Progression

### S1. Overview of the Modeling Framework

The supplementary methods are organized into two linked modeling components.

First, we considered a simplified natural history model that excludes transmission and demography and is used to fit cohort-derived progression data. This component was used to evaluate how alternative assumptions about the asymptomatic tuberculosis(aTB) dwell-time distribution, including the exponential case and Gamma formulations with larger shape parameters, affect the model's ability to reproduce observed progression patterns.

Second, we embedded the same aTB or subclinical progression structure within an extended population-level transmission model that incorporates a susceptible pool, transmission, demography, natural mortality, and TB-related mortality. This second component was used for calibration to population-level WHO incidence estimates for India and for the projection analyses reported in the main text.

For clarity, Part I below describes the simplified cohort-based progression model, and Part II describes the full transmission model used for population-level calibration.

### Part I. Simplified Natural History Model for Cohort Calibration

### S2. Model Structure for Cohort Calibration

We describe the simplified deterministic compartmental model used to fit cohort-derived progression data and to compare alternative assumptions about aTB disease progression.

Here, $I$ denotes early infection, $M$ minimal disease, $S_1, \ldots, S_k$ represent $k$ sequential subclinical/aTB compartments, $C$ clinical disease, $R$ recovered individuals, and $D$ cumulative deaths.

##### S2.1 System of Differential Equations

$$
\begin{aligned}
\frac{dI}{dt} &= -(\alpha+\beta+\gamma)\,I,\\
\frac{dM}{dt} &= \beta I - (\delta+\epsilon)\,M + \sum_{j=1}^{k}\zeta_j S_j,\\
\frac{dS_1}{dt} &= \gamma I + \epsilon M - (\kappa+\zeta_1)\,S_1 + \theta\,C,\\
\frac{dS_j}{dt} &= \kappa S_{j-1} - (\kappa+\zeta_j)\,S_j, \quad j=2,\dots,k-1,\\
\frac{dS_k}{dt} &= \kappa S_{k-1} - (\kappa+\zeta_k)\,S_k,\\
\frac{dC}{dt} &= \kappa S_k - (\theta+\mu)\,C,\\
\frac{dR}{dt} &= \alpha I + \delta M, \qquad \frac{dD}{dt} = \mu C.
\end{aligned}
\tag{1}
$$

##### S2.2 Model Diagram

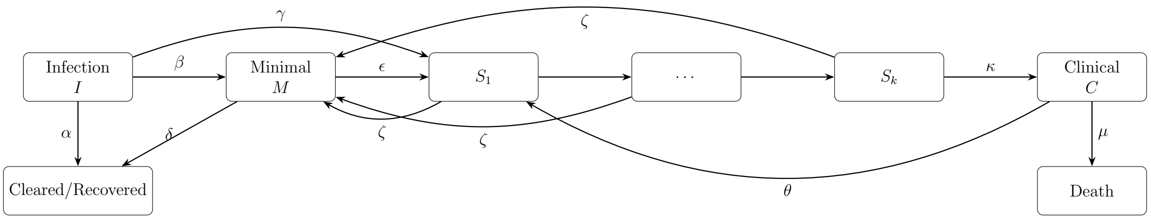


**Figure S1:** Uniform return from all $S_j$ to $M$.

### S3. Gamma Representation of aTB Progression

The aTB/subclinical phase is represented by $k$ sequential compartments with constant progression rate $\kappa$.

The implied dwell time distribution follows an Gamma distribution[Wearing et al., 2005]:

$$T \sim \text{Gamma}(k, \kappa),$$

with mean durationCox [1955]

$$\mathbb{E}[T] = \frac{k}{\kappa}.$$

For $k = 1$, the distribution reduces to the exponential case, corresponding to a constant hazard and memoryless progression.

Increasing $k$ reduces early transition probability and induces time-dependent hazards while preserving the same mean duration.

### S4. Derived Clinical Incidence Quantity from the Simplified Model

Although the simplified model is primarily used for fitting cohort-derived progression data rather than population-level incidence targets, it is useful to define the clinical progression flow in a form that is consistent with the full transmission model described in Part II.

Annual clinical incidence per $10^5$ population is defined as

$$\mathrm{Inc}_y = \frac{10^5}{\overline{N}_y} \int_y^{y+1} \kappa\, S_k(t)\, dt.$$

This quantity is reported here mainly to establish notation and to facilitate comparison with the population-level observation model introduced later.

### S5. Numerical Solution

The system of ordinary differential equations was solved numerically using a fourth–fifth order Runge–Kutta method (MATLAB `ode45`)Dormand and Prince [1980], The MathWorks Inc. [2024].

Relative and absolute tolerances were set to

$$\texttt{RelTol} = 10^{-7}, \qquad \texttt{AbsTol} = 10^{-9}.$$

All annual flow quantities were computed using trapezoidal numerical integration over 200 equally spaced time points within each yearly interval[Quarteroni et al., 2006].

### S6. Bayesian Calibration to Cohort-Derived Progression Data

Model calibration for the simplified natural history model was conducted within a Bayesian framework using Adaptive Metropolis Markov Chain Monte Carlo sampling.

Independent uniform priors were assigned to all parameters within biologically plausible bounds.

In addition to population-level incidence and mortality targets used in the full model, the simplified model component was calibrated to cohort-derived progression data.

Model fit was evaluated under different assumptions regarding the aTB dwell-time distribution ($k = 1, 50, 100$).

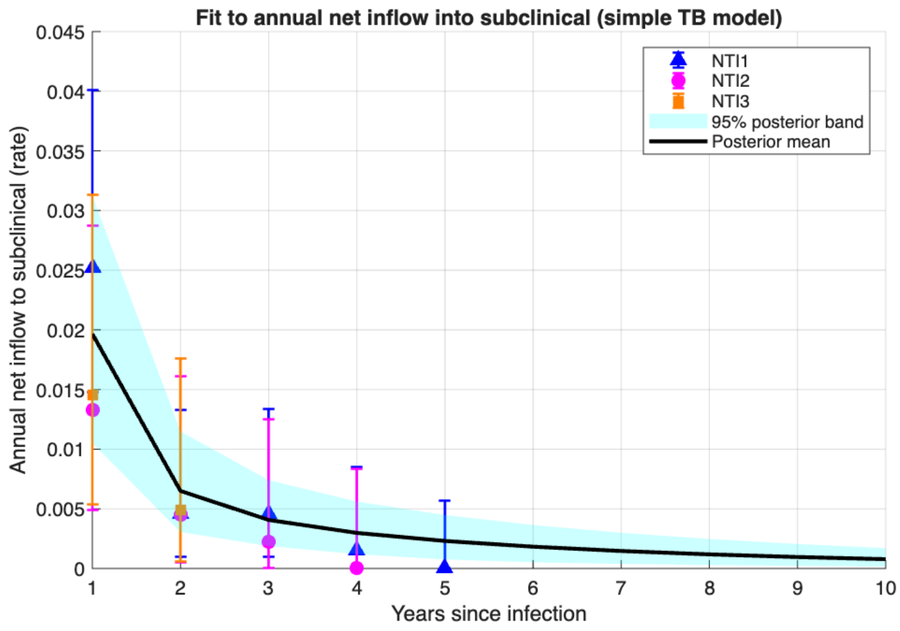


**(a)** Original scale.

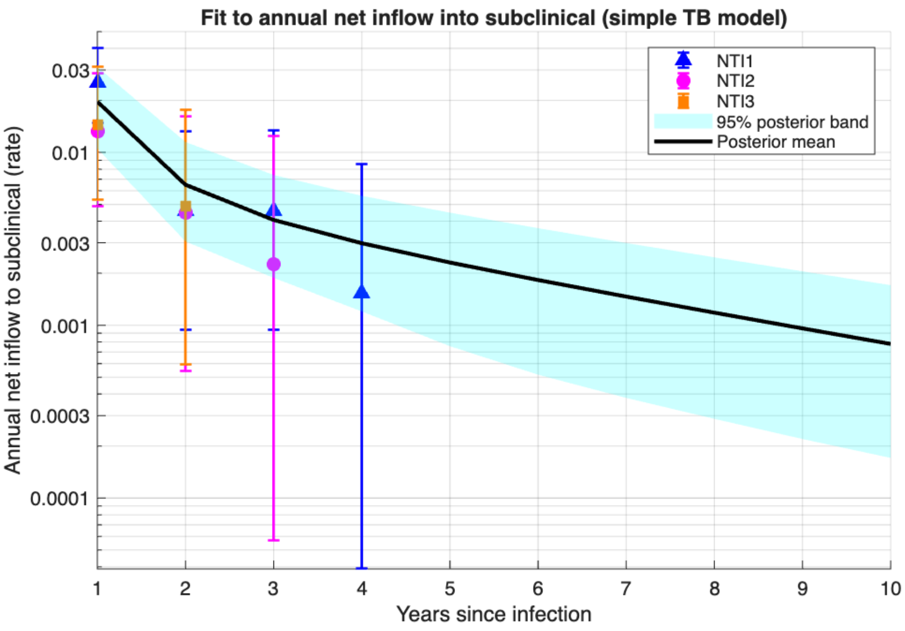


**(b)** Log scale.

**Figure S2:** Observed cohort progression data compared with posterior predictive median and 95% credible intervals under the exponential model ($k = 1$).

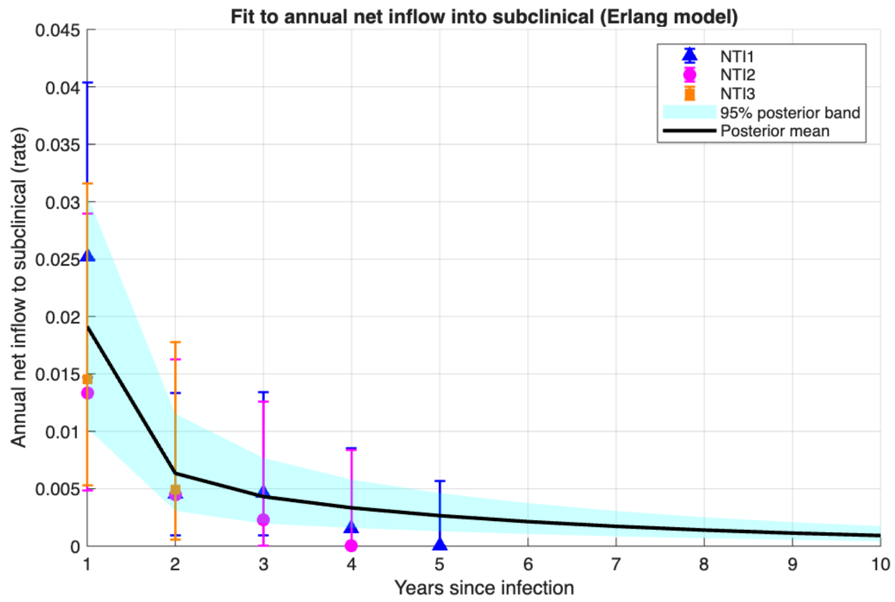


**(a)** Original scale.

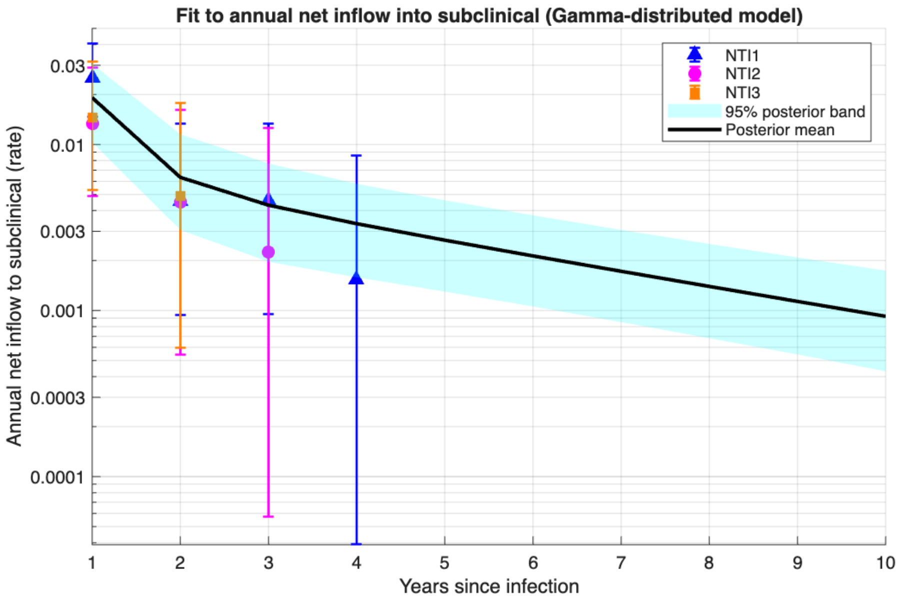


**(b)** Log scale.

**Figure S3:** Observed cohort progression data compared with posterior predictive median and 95% credible intervals under the Gamma-distributed model ($k = 50$).

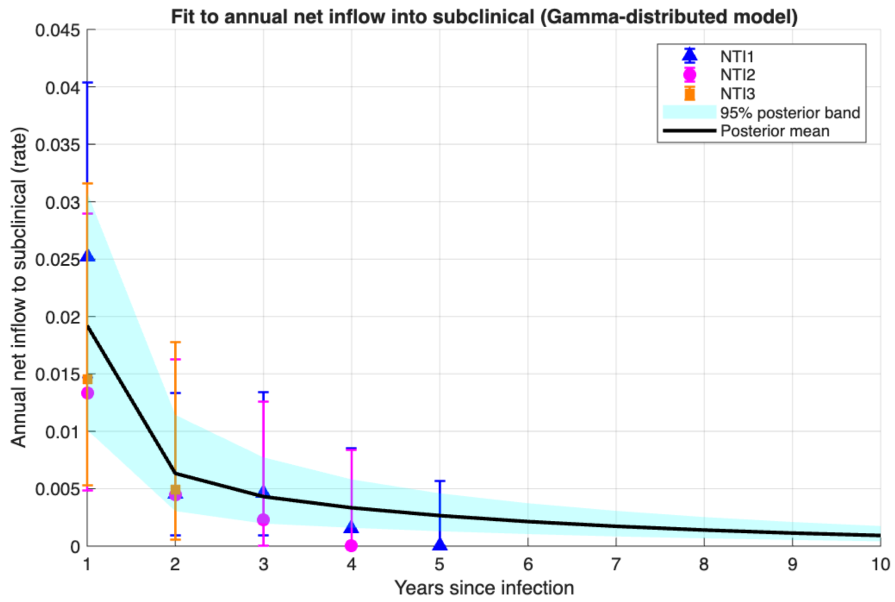


**(a)** Original scale.

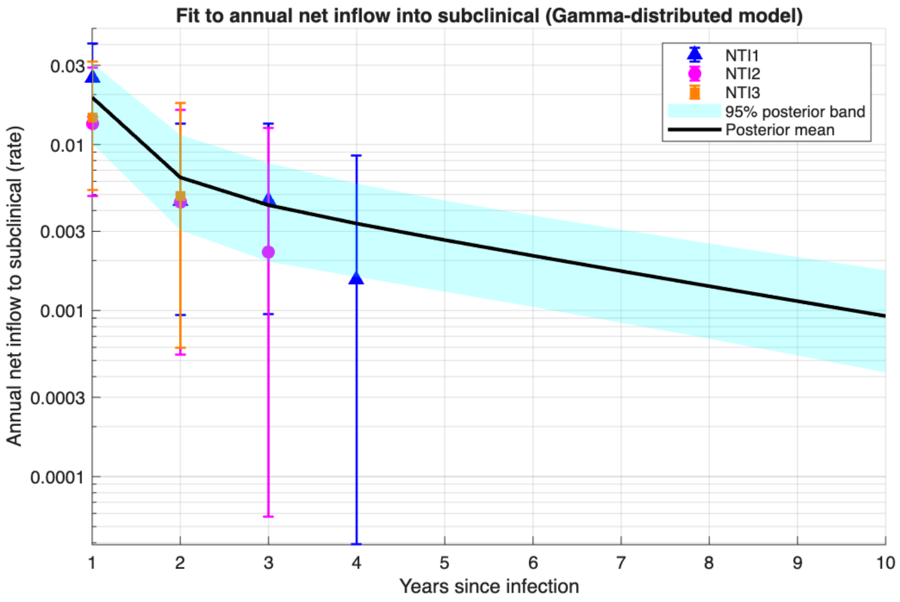


**(b)** Log scale.

**Figure S4:** Observed cohort progression data compared with posterior predictive median and 95% credible intervals under the Gamma-distributed model ($k = 100$).

### Part II. Extended Transmission Model for Population-Level Calibration

#### S7. Extended Transmission Model with Demography

To calibrate the model to population-level WHO estimates for India[World Health Organization, 2025], we extended the cohort-fitting model to incorporate a susceptible pool, time-varying demography, natural mortality, and TB-excess mortality. Model state variables represent proportions (or equivalently, densities scaled by an arbitrary reference population), so annual incidence per $10^5$ is computed from integrated flow rates multiplied by $10^5$.

#### S7.1 Model Compartments

$S_0$ denotes susceptible individuals, $I$ early infection, $M$ minimal disease, $S_1, \ldots, S_k$ the sequential subclinical compartments, $C$ clinical disease, $R$ recovered, and $D$ cumulative deaths.

#### S7.2 Force of Infection

Transmission is driven by clinical disease, subclinical disease, and minimal disease:

$$\lambda(t) \;=\; \beta_{\text{trans}} \frac{C(t) + \rho_S \sum_{j=1}^{k} S_j(t) + \rho_M M(t)}{N(t)},$$

where $N(t)$ is the total living population and $\rho_S, \rho_M$ scale relative infectiousness.

#### S7.3 Demography

Let $\nu$ denote the natural mortality rate and $g(t)$ the time-varying per-capita population growth rate derived from WHO annual population estimates (Section S7.6). Births are specified as

$$\Pi(t) \;=\; \big(g(t) + \nu\big)\, N(t),$$

so that in the absence of TB-excess mortality, the living population grows approximately at rate $g(t)$.

The total living population is

$$N(t) \;=\; S_0 + I + M + \sum_{j=1}^{k} S_j + C + R.$$

#### S7.4 System of Differential Equations

The extended model is given by

$$
\begin{aligned}
\frac{dS_0}{dt} &= \Pi(t) - \lambda(t)\, S_0 - \nu S_0, \\
\frac{dI}{dt} &= \lambda(t)\, S_0 - (\alpha + \beta + \gamma + \nu)\, I, \\
\frac{dM}{dt} &= \beta I - (\delta + \epsilon + \nu + \mu_M)\, M + \zeta \sum_{j=1}^{k} S_j, \\
\frac{dS_1}{dt} &= \gamma I + \epsilon M - (\kappa + \zeta + \nu + \mu_S)\, S_1 + \theta C, \\
\frac{dS_j}{dt} &= \kappa S_{j-1} - (\kappa + \zeta + \nu + \mu_S)\, S_j, \qquad j = 2, \ldots, k, \\
\frac{dC}{dt} &= \kappa S_k - (\theta + \nu + \mu_C)\, C, \\
\frac{dR}{dt} &= \alpha I + \delta M - \nu R, \\
\frac{dD}{dt} &= \nu N(t) + \mu_C C + \mu_M M + \mu_S \sum_{j=1}^{k} S_j. 
\end{aligned} \tag{2}
$$

For $k = 1$, the subclinical phase reduces to a single compartment (the exponential case).

#### S7.5 Observation Model: Annual Clinical Incidence (WHO Targets)

Annual clinical incidence per $10^5$ population in calendar year $y$ is computed as the integrated flow into $C$ over the yearly interval:

$$\text{Inc}_y \;=\; 10^5 \int_y^{y+1} \kappa\, S_k(t)\, dt.$$

For the exponential ($k = 1$) model, incidence is computed as

$$\text{Inc}_y \;=\; 10^5 \int_y^{y+1} \eta\, S_1(t)\, dt.$$

Model outputs were compared to WHO India clinical incidence estimates for 2010–2023.

#### S7.6 Population Input and Growth-Rate Construction

Worldometer Population estimates were entered as year-specific anchor points and interpolated to obtain an annual population series $N_y$ for 1990–2024[Worldometer, n.d.]. The implied annual continuous growth rate was computed as

$$g_y \;=\; \log\!\left(\frac{N_{y+1}}{N_y}\right),$$

and treated as piecewise constant within each calendar year.

#### S7.7 Numerical Solution

The extended population-level model was solved numerically using the same fourth–fifth order Runge–Kutta method (MATLAB `ode45`) described in Part IDormand and Prince [1980], The MathWorks Inc. [2024]. Relative and absolute tolerances were again set to

$$\texttt{RelTol} = 10^{-7}, \qquad \texttt{AbsTol} = 10^{-9}.$$

All annual flow quantities were computed using trapezoidal numerical integration over 200 equally spaced time points within each yearly interval[Quarteroni et al., 2006].

#### S7.8 Bayesian Calibration to WHO Incidence

Calibration was performed in a Bayesian framework using Adaptive Metropolis MCMC[Hastings, 1970, Andrieu and Thoms, 2008, Ryckman et al., 2022]. Independent uniform priors were assigned to all parameters within prespecified bounds.

Let $\mathbf{y}$ denote the WHO annual incidence observations and $\boldsymbol{\mu}(\theta)$ the corresponding model-predicted annual incidence. We assumed a Gaussian observation model

$$y_i \sim \mathcal{N}\left(\mu_i(\theta),\ \sigma_i^2 + \sigma_{\text{add}}^2\right),$$

where $\sigma_i$ was derived from reported 95% uncertainty intervals and $\sigma_{\text{add}}$ is an additional error term to account for residual model discrepancy.

### 1 Further results

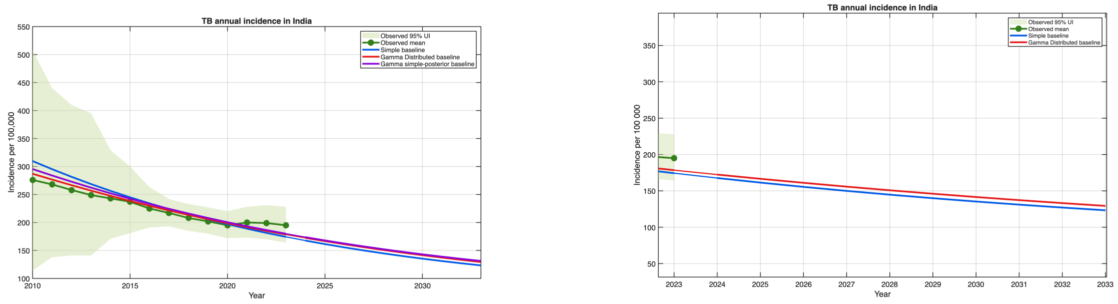


**(a)** Full trajectory (2010–2033) **(b)** Zoomed projection (2023–2033)

**Figure S5:** Posterior baseline incidence trajectories under alternative model structures. Green points and shaded area show WHO estimated incidence (mean and 95% uncertainty interval). Lines represent posterior predictive medians under the simple exponential model (blue), Gamma-distributed model (red), and Gamma-distributed model using the simple-model posterior (purple).

Beyond their impact on mean trajectories, distributional assumptions also affect parameter identifiability and the propagation of uncertainty. Simpler exponential models often allow different parameter combinations to fit historical data equally well, yet produce divergent future projections. By contrast, multi-stage formulations impose additional structural constraints on progression timing, which can reduce parameter trade-offs and yield more stable posterior predictive behavior. As a result, differences in uncertainty growth across model structures should be interpreted as reflecting structural assumptions rather than numerical artefacts.

W Keith Hastings. Monte carlo sampling methods using markov chains and their applications. 1970.

Alfio Quarteroni, Riccardo Sacco, and Fausto Saleri. *Numerical mathematics*, volume 37. Springer Science & Business Media, 2006.

Theresa S Ryckman, David W Dowdy, and Emily A Kendall. Infectious and clinical tuberculosis trajectories: Bayesian modeling with case finding implications. *Proceedings of the National Academy of Sciences*, 119(52):e2211045119, 2022.

The MathWorks Inc. *MATLAB R2024a*. The MathWorks Inc., Natick, Massachusetts, United States, 2024.

Helen J Wearing, Pejman Rohani, and Matt J Keeling. Appropriate models for the management of infectious diseases. *PLoS medicine*, 2(7):e174, 2005.

World Health Organization. Global tuberculosis report 2025: Tb profiles (india). `https://worldhealthorg.shinyapps.io/tb_profiles/?_inputs_&tab=%22charts%22&lan=%22EN%22&iso3=%22IND%22&entity_type=%22country%22`, 2025. [cited 2026-01-12].

Worldometer. India population (live) — worldometer. `https://www.worldometers.info/world-population/india-population/`, n.d. Accessed: 2026-02-28.

Table 1: Uniform prior bounds for model parameters in the Simple and Gamma-distributed formulations.

| Parameter | Lower bound | Upper bound |
|---|---|---|
| *Simple model* | | |
| $b_{\text{trans}}$ | 0.05 | 4.00 |
| $\rho_S$ | 0.00 | 1.00 |
| $\rho_M$ | 0.00 | 1.00 |
| $\alpha$ | 0.15 | 5.85 |
| $\beta$ | 0.08 | 2.93 |
| $\gamma$ | 0.08 | 2.93 |
| $\delta$ | 0.08 | 2.93 |
| $\epsilon$ | 0.08 | 2.93 |
| $\zeta$ | 0.08 | 2.93 |
| $\eta$ | 0.08 | 2.93 |
| $\theta$ | 0.08 | 2.93 |
| $\mu_C$ | 0.10 | 1.00 |
| $\mu_M$ | 0.00 | 0.20 |
| $\mu_S$ | 0.00 | 0.20 |
| $\nu$ | 0.005 | 0.030 |
| *Gamma-distributed model (shape parameter $k$)* | | |
| $b_{\text{trans}}$ | 0.05 | 4.00 |
| $\rho_S$ | 0.00 | 1.00 |
| $\rho_M$ | 0.00 | 1.00 |
| $\alpha$ | 0.15 | 5.85 |
| $\beta$ | 0.08 | 2.93 |
| $\gamma$ | 0.08 | 2.93 |
| $\delta$ | 0.08 | 2.93 |
| $\epsilon$ | 0.08 | 2.93 |
| $\kappa$ | $3k/2$ | $4k$ |
| $\zeta$ | 0.08 | 2.93 |
| $\theta$ | 0.08 | 2.93 |
| $\mu_C$ | 0.10 | 1.00 |
| $\mu_M$ | 0.00 | 0.20 |
| $\mu_S$ | 0.00 | 0.20 |
| $\nu$ | 0.005 | 0.030 |
